# A 3D CNN Classification Model for Accurate Diagnosis of Coronavirus Disease 2019 using Computed Tomography Images

**DOI:** 10.1101/2021.01.21.21249999

**Authors:** Yifan Li, Xuan Pei, Yandong Guo

## Abstract

The coronavirus disease (COVID-19) has been spreading rapidly around the world. As of August 25, 2020, 23.719 million people have been infected in many countries. The cumulative death toll exceeds 812,000. Early detection of COVID-19 is essential to provide patients with appropriate medical care and protect uninfected people. Leveraging a large computed tomography (CT) database from 1,112 patients provided by China Consortium of Chest CT Image Investigation (CC-CCII), we investigated multiple solutions in detecting COVID-19 and distinguished it from other common pneumonia (CP) and normal controls. We also compared the performance of different models for complete and segmented CT slices. In particular, we studied the effects of CT-superimposition depths into volumes on the performance of our models. The results show that the optimal model can identify the COVID-19 slices with 99.76% accuracy (99.96% recall, 99.35% precision and 99.65% F1-score). The overall performance for three-way classification obtained 99.24% accuracy and the area under the receiver operating characteristic curve (AUROC) of 0.9986. To the best of our knowledge, our method achieves the highest accuracy and recall with the largest public available COVID-19 CT dataset. Our model can help radiologists and physicians perform rapid diagnosis, especially when the healthcare system is overloaded.

## Introduction

The outbreak of the 2019 novel coronavirus (SARS-CoV-2) began in early December of 2019 (Munster et. al. 2020; Wang et. al. 2020). The infection has an average incubation period of 5.2 days and can cause fever, cough and other flu-like symptoms. It can affect multiple tissues and organ systems, and diseases caused by viruses are collectively referred to as coronavirus disease 2019 (COVID-19). Many infected patients develop pneumonia and rapidly, severe acute respiratory failure, with very poor prognosis and high mortality (Guan et. al. 2020; Huang et. al. 2020). Person-to-person transmission has been confirmed (Chan et. al. 2020; Phan et. al. 2020; Rothe et. al. 2020; Zhu et. al. 2020). Compared with the prior Severe Acute Respiratory Syndrome (SARS) and Middle East Respiratory Syndrome (MERS), although COVID-19 has a relatively lower fatality rate, it has spread to more places and caused more deaths (Wu and McGoogan 2020; Mahase 2020). The World Health Organization (WHO) declared COVID-19 to be pandemic. Therefore, it is necessary to build an accurate diagnostic solution for early intervention and close monitoring of such a pandemic which could facilitate slowing the spread of the virus and contain the disease.

In clinics, a positive molecular polymerase chain reaction (PCR) test is the golden standard to make a definitive diagnosis of COVID-19 infection (Zu et. al. 2020). However, the high false negative rate (Chan et. al. 2020) and unavailability of PCR assay in the early stage of an outbreak may delay the identification of potential patients (Ouyang et. al. 2020). Chest computed tomography (CT) is an important tool to diagnose lung diseases including pneumonia. CT scan procedures have a faster turnaround time than molecular diagnostic tests performed in standard laboratories and can provide more detailed pathological information. For example, almost all COVID-19 patients have some typical radiographic features in chest CT, including ground-glass opacities (GGO), multifocal patchy consolidation, and/or interstitial changes with a peripheral distribution (Chung et. al. 2020). Therefore, chest CT has been recommended as a major tool for clinical diagnosis especially in the hard-hit region such as Hubei province, China (Zu et. al. 2020). Since seasonal influenza can also cause viral pneumonia, it is also crucial to distinguish COVID-19 from common influenza or other types of pneumonia such as viral and bacterial pneumonia. Considering the high demand for chest CT screening and the workload of radiologists, especially as an outbreak occurs, we design a deep-learning method using CT images to classify COVID-19, common pneumonia (CP), and normal controls.

In recent years, the application of deep learning in many medical fields has made exciting novel progresses (Chilamkurthy et. al. 2018; Esteva et. al. 2019; Li et. al. 2018; Norgeot et. al. 2019; Poplin et. al. 2018; Ting et. al. 2017; Topol 2019), stimulating the development and innovation of new radiological diagnostic technology. With the outbreak of epidemic, deep-learning methods were used in the diagnosis, prognosis, detection, and treatment of COVID-19. Butt et. al. (2020) compared multiple convolutional neural network (CNN) models to classify CT samples with COVID-19, influenza viral pneumonia, or no-infection and calculated a sensitivity of 98.2% and a specificity of 92.2%. Ouyang et al. (2020) developed a dual-sampling attention network to automatically diagnose COVID-19 from CP in chest CT, and calculated an accuracy of 87.5%. Zhang et. al. (2020) developed an artificial intelligence (AI) system that was able to diagnose COVID-19 and provide accurate clinical prognosis.

As a summary, the contributions of our work are in three-fold:

1. We investigated several 3D CNN technologies, including basic block, bottleneck block and (2+1)D convolution, and reported the optimal solution to detect COVID-19 from CT images.
2. We used different depths to superimpose CT slices for preprocessing to obtain more information between CT slices. The superimposed volume was used as the input of the 3D classification network. The experimental results demonstrate that the depth of volume has a great influence on the model effect.
3. We conducted experiments with a large CT dataset provided by the China Consortium of Chest CT Image Investigation (Zhang et. al. 2020) including complete CT slices and segmented CT slices. Experimental results demonstrate our method can identify the COVID-19 slices with 99.76% accuracy, 99.96% recall, 99.35% precision and 99.65% F1-sorce. The overall performance for three-way classification obtained 99.24% accuracy and the area under the receiver operating characteristic curve (AUROC) of 0.9986. To the best of our knowledge, this is the most accurate result with the largest public available dataset.

## Methodology

### Data Set

A large CT dataset from the China Consortium of Chest CT Image Investigation (CC-CCII) was used (Zhang et. al. 2020), which consists of a total of 137,256 complete CT images from 691 patients and 42,861 segmented CT images from 421 patients (Figure 1).

**Figure 1.**
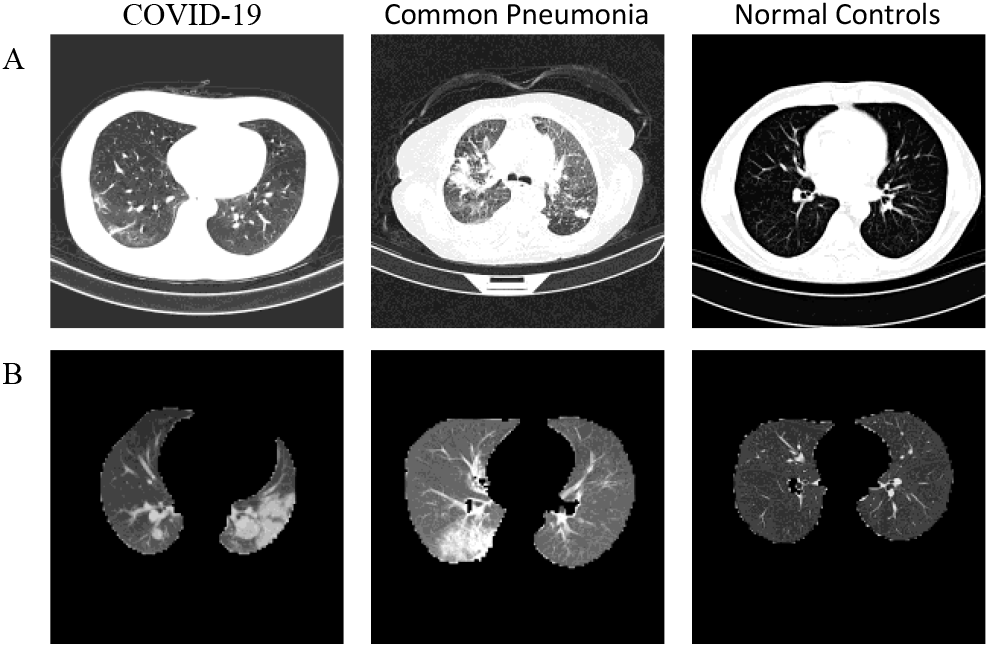
Typical transverse-section CT images: (A), complete CT images; row, segmented CT images.

A total of 110,420 complete CT images (80.4%) were employed to train and valid our model for discriminating COVID-19 from other CP and normal controls (Table 1). The remaining 26,836 CT images (19.6%) were used as the test set. In addition, the test set used CT slices selected from the people who were not included in the training and validation stages. Viral pneumonia, bacterial pneumonia and mycoplasma pneumonia are included in CP group, all of which are the most common causes of pneumonia in China. We also tried to use segmented CT images to train, valid and test our model (Table 2).

**Table 1.**
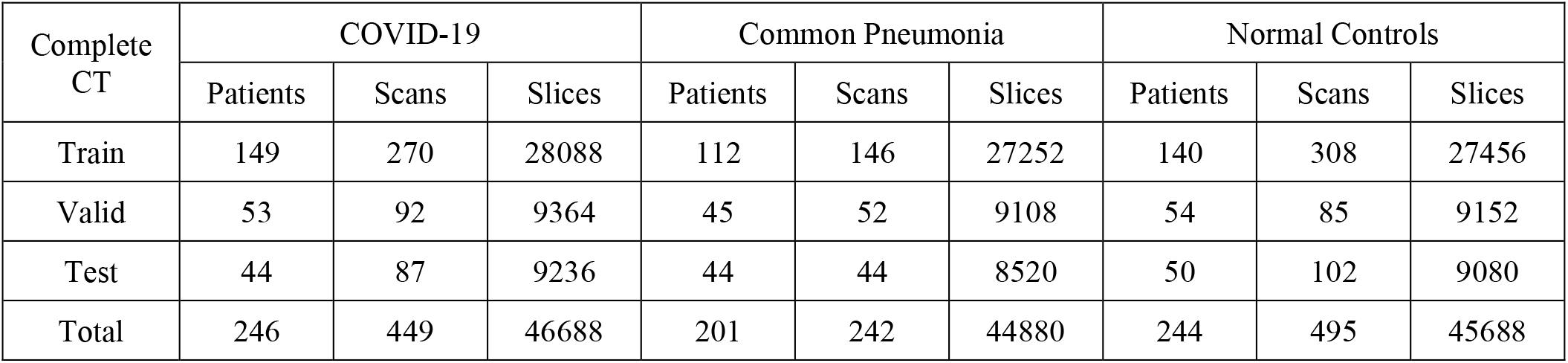
The complete CT dataset of characteristics in identifying COVID-19 from other CP and normal controls.

| Complete CT | COVID-19 |  |  | Common Pneumonia |  |  | Normal Controls |  |  |
| --- | --- | --- | --- | --- | --- | --- | --- | --- | --- |
|  | Patients | Scans | Slices | Patients | Scans | Slices | Patients | Scans | Slices |
| Train | 149 | 270 | 28088 | 112 | 146 | 27252 | 140 | 308 | 27456 |
| Valid | 53 | 92 | 9364 | 45 | 52 | 9108 | 54 | 85 | 9152 |
| Test | 44 | 87 | 9236 | 44 | 44 | 8520 | 50 | 102 | 9080 |
| Total | 246 | 449 | 46688 | 201 | 242 | 44880 | 244 | 495 | 45688 |

**Table 2.** The segmented CT dataset of characteristics in identifying COVID-19 from other CP and normal controls

| Segmented CT | COVID-19 |  |  | Common Pneumonia |  |  | Normal Controls |  |  |
| --- | --- | --- | --- | --- | --- | --- | --- | --- | --- |
|  | Patients | Scans | Slices | Patients | Scans | Slices | Patients | Scans | Slices |
| Train | 79 | 98 | 11648 | 2 | 3 | 222 | 175 | 175 | 14144 |
| Valid | 26 | 26 | 3904 | 1 | 1 | 77 | 56 | 56 | 4736 |
| Test | 25 | 32 | 3456 | 1 | 1 | 66 | 56 | 56 | 4608 |
| Total | 130 | 156 | 19008 | 4 | 5 | 365 | 287 | 287 | 23488 |

### Preprocess

CT slices were normalized to 512 × 152 × 3 for the height, width and channel, respectively. In order to leverage the 3D volume of CT images to capture a wide range of spatial information both within the CT slices and between CT slices (Li et. al. 2020), we stacked *n* CT slices in a CT scan vertically to form a volume, where *n* denotes the depth. We then transposed the volume from *D* × *H* × *W* × *C* to *C* × *D* × *H* × *W*, deriving a tensor. The diagnostic classifier took the tensor as input, and used the classification network to generate the three-level probabilities of COVID-19, CP and normal controls and produce the class with the maximum probability after a softmax activation function.

### Network Architecture

The detailed structure of the three-way classification network was shown in Table 3, based on 3D ResNet-18 (Hara et. al. 2017). The network used multiple 3D basic blocks with residual connections which could continuously extract local and global contextual features, and used a fully connected layer followed by a softmax activation function to calculate final predictions. For the three types of diagnostic results, including COVID-19, CP and normal controls, the model yielded the class with the maximum probability.

**Table 3.** Network architectures. Each convolutional layer is followed by batch normalization (Ioffe and Szegedy 2015) and a ReLU (Nair and Hinton 2010). Down-sampling is performed by conv3_1, conv4_1, and conv5_1 with a stride of 2. *F* is the number of feature channels corresponding in Figure 2, and *N* is the number of blocks in each layer.

| Layer Name | Architecture |  |  |  |  |  |
| --- | --- | --- | --- | --- | --- | --- |
|  | 18-layer |  | 34-layer |  | 50-layer |  |
|  | F | N | F | N | F | N |
| Conv1 | $7 \times 7 \times 7$, 64, stride 1 (D), 2 (HW) | | | | | |
| Conv2 | $3 \times 3 \times 3$ max pool, stride 2 | | | | | |
|  | 64 | 2 | 64 | 3 | 64 | 3 |
| Conv3 | 128 | 2 | 128 | 4 | 128 | 4 |
| Conv4 | 256 | 2 | 256 | 6 | 256 | 6 |
| Conv5 | 512 | 2 | 512 | 3 | 512 | 3 |
| Global average pool, fully-connected, softmax |  |  |  |  |  |  |
| Block | Basic |  | Basic |  | Bottleneck |  |

We employed cross entropy loss between the final predictions and ground truth labels to train the 3D classification network. The Adam optimizer with an initial learning rate at 0.001 was used in training. The learning rate decays by a factor of 0.1 for every 10 epochs. The training epoch is 20 in total. The training batch size is given in the next section. The whole training, validation and testing procedures were conducted with Pytorch (v.1.2.0) on NVIDIA Tesla V100 SXM2 graphical processing units (Paszke et. al. 2019).

### Basic Block

The basic block of ResNets consists of two convolutional layers (Figure 2). After each convolutional layer there are batch normalization and ReLU. A shortcut pass connects the top of the block to the layer just before the last ReLU in the block. ResNet-18 and 34 adopt the basic blocks. We use identity connections and zero padding as the shortcuts to the basic blocks to avoid increasing the number of parameters of these relatively shallow networks (He et. al. 2016).

**Figure 2.**
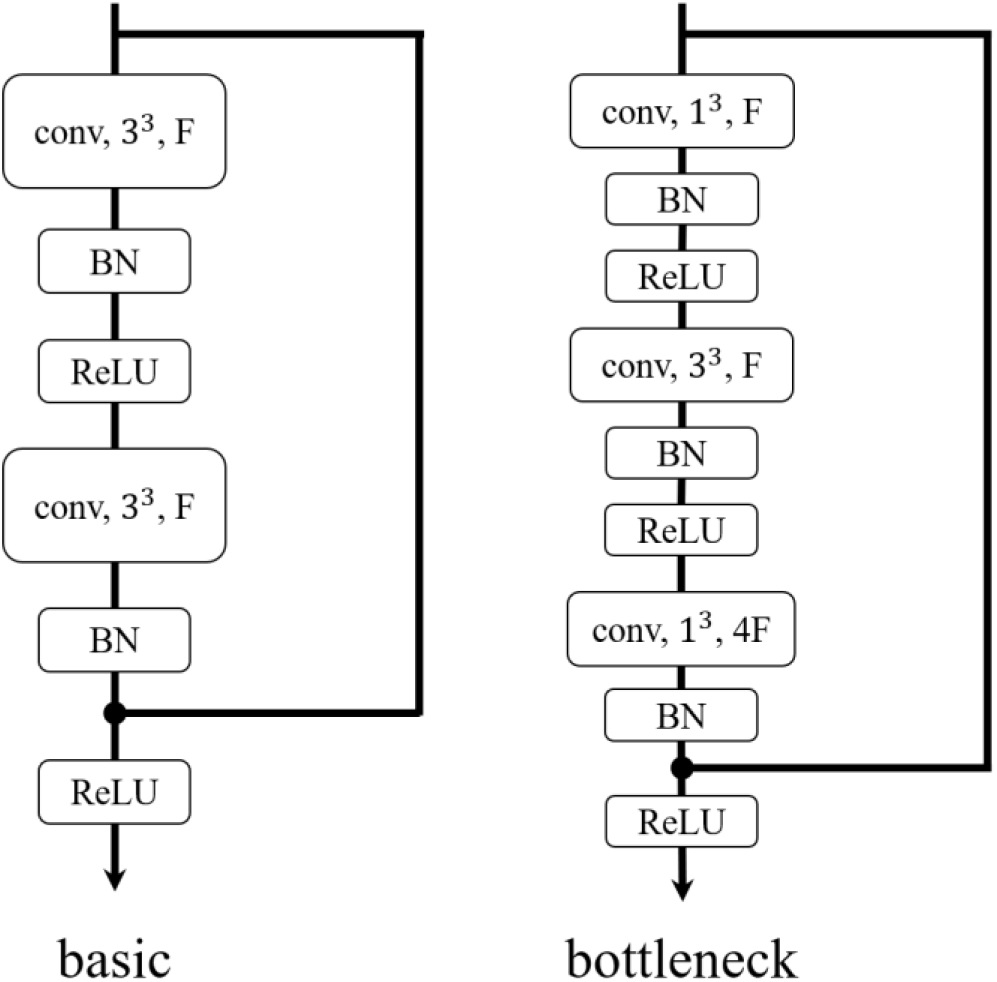
Blocks of architectures. *x*^3^ denotes the kernel size, and *F* denotes the number of feature channels.

### Bottleneck Block

The bottleneck block of ResNets consists of three convolutional layers (Figure 2). The kernel sizes of the first and third convolutional layers are 1 × 1 × 1, and the second convolutional is 3× 3 × 3. Each convolutional layer is followed by batch normalization and ReLU. The shortcut pass of this block is the same as the basic block. ResNet-50, 101, 152, and 200 all adopt the bottleneck block. We use identity connections except for those that are used to increase the dimensions (He et. al. 2016).

### (2+1)D Convolutions

In order to decompose spatial and temporal modeling into two separate steps, Tran et. al. (2018) thus designed a network architecture named R(2+1)D, where they replaced the 3D convolutional filters of size *N* × *t* × *d* × *d* with a (2+1)D block consisting of 2D convolutional filters of size *N* × 1 × *d* × *d* and temporal convolutional filters of size *N* × *t* × 1 × 1 (Figure 3).

**Figure 3.**
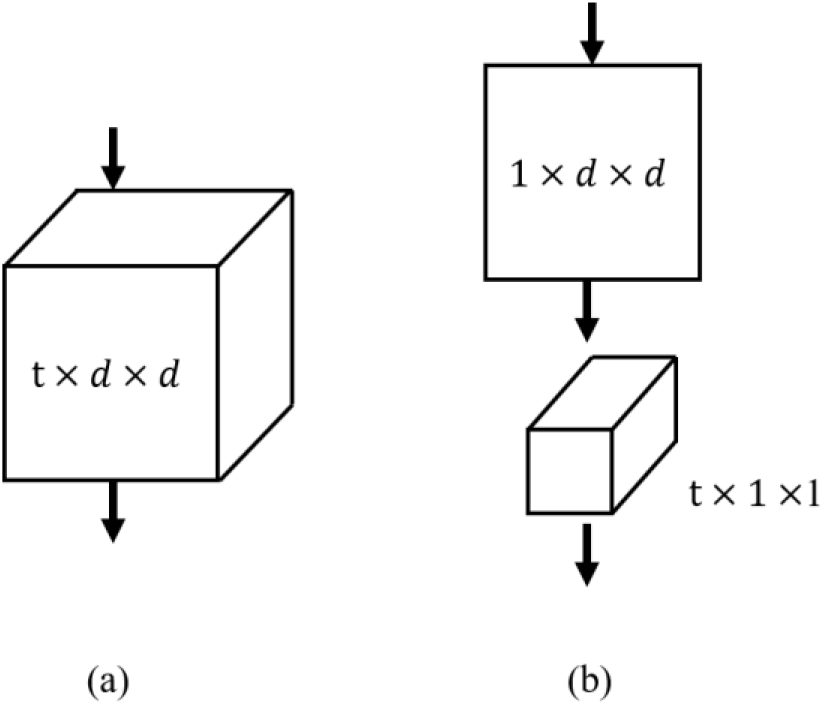
3D vs (2+1)D Convolution. (a) Full 3D convolution. (b) (2+1)D convolution.

The first advantage is that a nonlinear correction is added between these two operations. Compared with a full 3D convolutional network using the same number of parameters, this effectively doubles the number of nonlinearities, allowing the model to represent more complex functions. The second potential benefit is that decomposition helps to optimize, resulting in lower training loss and lower test loss in practice.

### Classification Performance Analysis

The accuracy of a method determines the correctness of the predicted value, the precision determines the repeatability of the measurement or the correctness of the predicted value, and the recall or sensitivity indicates how many of correct results are discovered. The F1-score is used as an overall measure of the model accuracy, combining precision and recall measures to calculate a balanced average result. The formulas for these values are summarized as follows, where TP, TN, FP and FN are true positive, true negative, false positive, and false negative respectively.

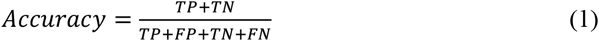

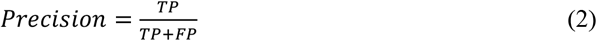

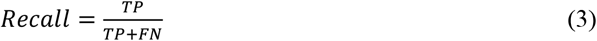

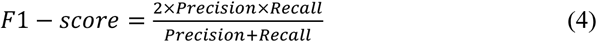

## Experimental Results

### Complete and Segmented CT Slices

Zhang et al (2020) employed a diagnostic system based on a lung-lesion segmentation model. The diagnosis classification took the segmented CT slices as an input generated by segmentation networks where the depth and batch size are 64 and 8 respectively. The CC-CCII dataset contains both complete and segmented CT slices. Due to the small number of segmented CT slices, we used the same number of complete CT slices for training, validation and test (Table 2).

The accuracy is used to evaluate the overall performance for three-way classification. With the same number of slices, the accuracy of complete CT is 5.5% higher than that of segmented and the recall is 9.2% higher (Table 4). Therefore, complete CT slices were used for the rest of this study. For complete CT slices, a smaller data set achieved higher accuracy because all 42,861 slices contained only 365 CP slices. This made the three-way classification model close to the binary classification model.

**Table 4.** Accuracy and recall of complete and segmented CT Images.

| Depth and batch size | Slices | Accuracy | Recall |
| --- | --- | --- | --- |
| 64 8<br>(complete) | 137,256 | 0.929612 | 0.907801 |
| 64 8<br>(complete, same number) | 42,861 | 0.981651 | 0.981481 |
| 64 8<br>(segmented) | 42,861 | 0.926606 | 0.888889 |

### Depth and Batch Size

The depth and batch size have strong influence on the model training effect and final accuracy. We first experimented with the three-way classification effect of different depths when batch size was equal to 8 (Table 5 and Figure 4). The data set settings are shown in Table 1.

**Table 5.** The effect of depths on accuracy and recall.

| Depth | Batch size | Accuracy | Recall |
| --- | --- | --- | --- |
| 2 | 8 | 0.848201 | 0.979645 |
| 4 | 8 | 0.914688 | 0.940234 |
| 8 | 8 | 0.968396 | 0.950355 |
| 16 | 8 | 0.967273 | 0.948007 |
| 32 | 8 | 0.963604 | 0.934142 |
| 64 | 8 | 0.929612 | 0.907801 |

**Figure 4.**
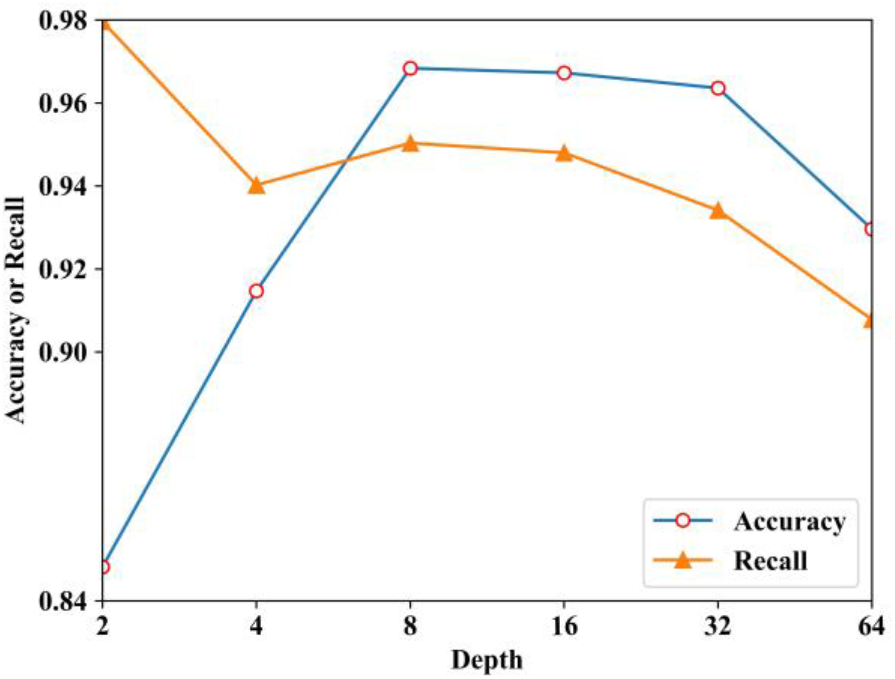
The effect of depths on accuracy and recall.

Due to the small batch size, it was difficult for the model to converge when the depth was 2 or 4, so we only compared the depth from 8 to 64. From Table 5 and Figure 4 we can conclude that the accuracy and recall increase as the depth decreases under the same batch size. We next experimented with the effect of different batch sizes on the performance of the three-classification model (Table 6 and Figure 5).

**Table 6.**
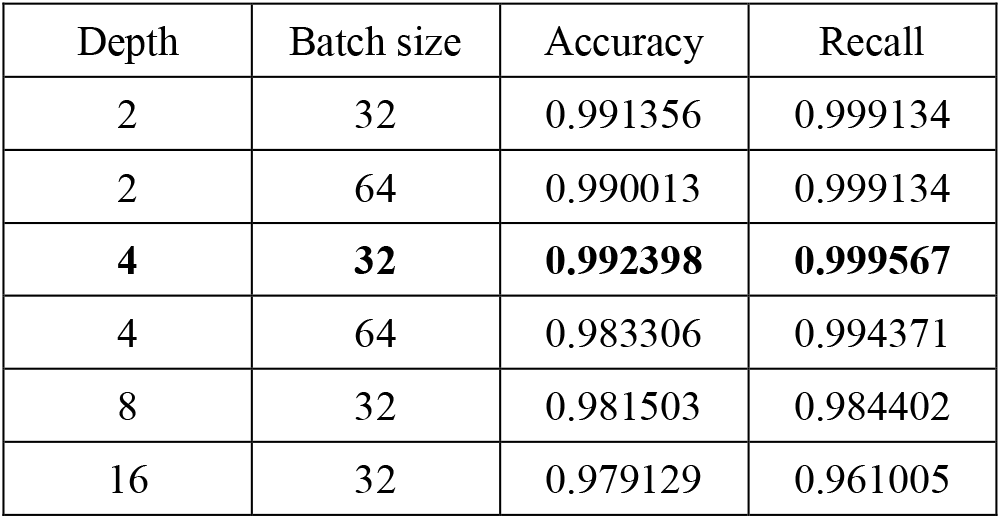
The effect of batch sizes on the accuracy and recall

**Table 7.** Comparison of classification results using different models.

| Model | Accuracy | Recall |
| --- | --- | --- |
| ResNet-18 | 0.992398 | 0.999567 |
| ResNet-18(2+1)D | 0.988523 | 0.997835 |
| ResNet-34 | 0.980027 | 0.971416 |
| ResNet-34(2+1)D | 0.971829 | 0.963188 |
| ResNet-50 | 0.980176 | 0.972282 |

**Figure 5.**
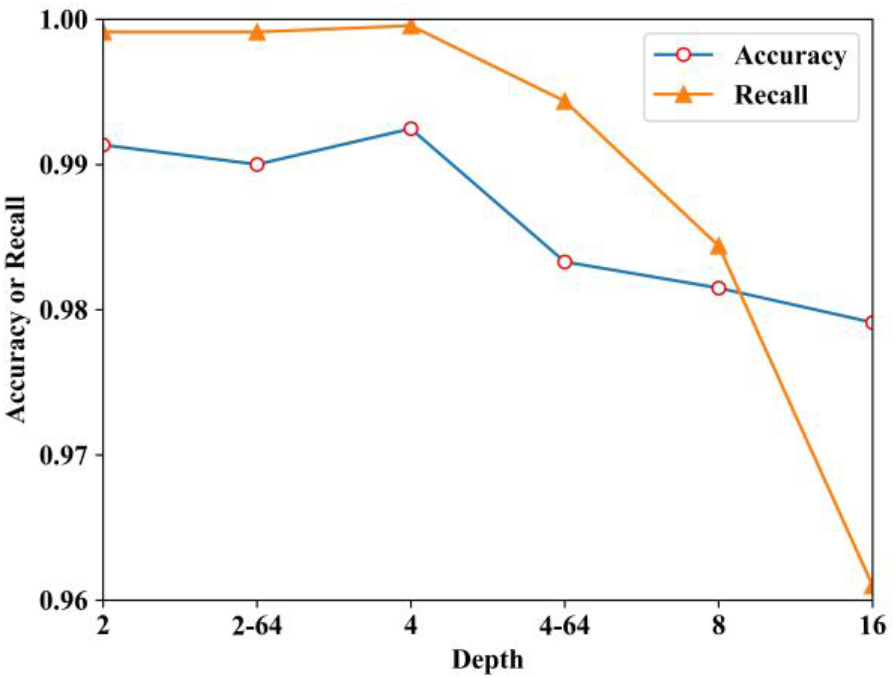
The effect of batch sizes on the accuracy and recall.

From Table 6 and Figure 5 we conclude that the accuracy and recall are significantly improved as the batch size is increased to 32. Especially when the depth is 2 and 4, the accuracy and recall have reached 0.99, but increasing the batch size to 64 does not further improve the model performance and the accuracy and recall rate even decrease.

### Different Models

We use optimal depth 4 and a batch size of 32 to train different models, including 3D ResNet-18, 3D ResNet-34, 3D ResNet-50, (2+1)D ResNet-18.

As the number of ResNet layers deepened to 34, the network appeared over-fitting and the accuracy and recall dropped slightly. The performance of ResNet-34 and Res-Net-50 were relatively close. After replacing the 3D convolution with (2+1)D, the accuracy and recall both decreased.

Our optimal model was able to discriminate COVID-19 from other two classes (other CP and normal controls) with 99.76% accuracy, 99.96% recall, 99.35% precision and 99.65% F1-sorce (Figure 6). The overall performance for three-way classification obtained 99.24% accuracy and an AUROC of 0.9986.

**Figure 6.**
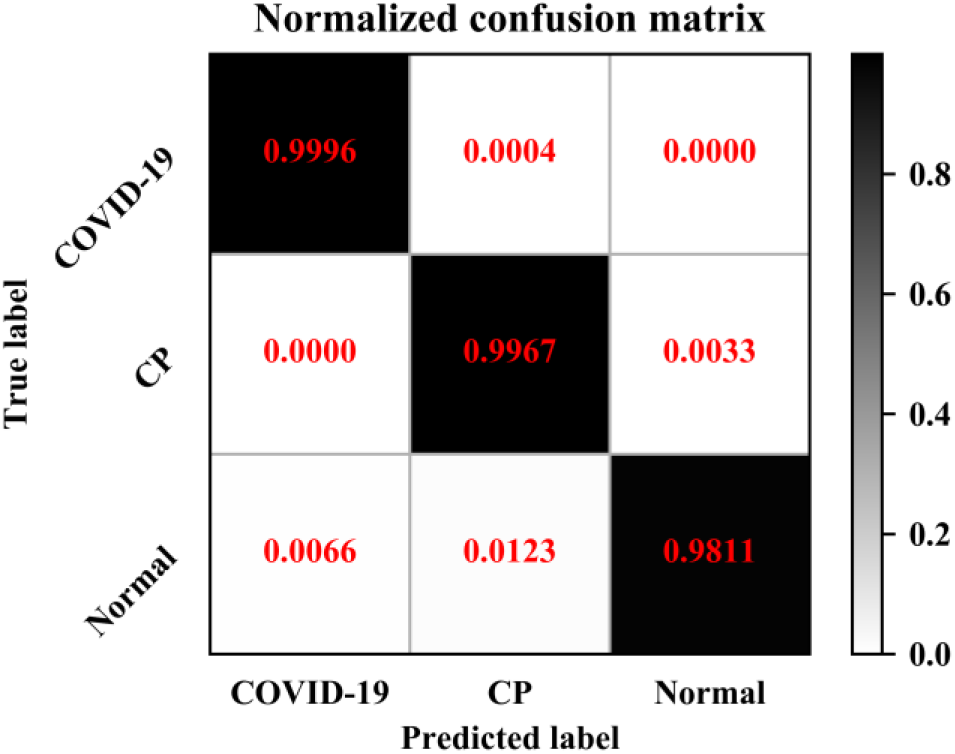
Normalized confusion matrix of depth 4 and batch size 32. The model used 3D ResNet-18.

## Discussion

For COVID-19, getting a diagnosis as soon as possible is essential. PCR is the gold standard for diagnosing COVID-19, but it takes a few days to get the final results and the testing capacity in many places is limited, especially in the early outbreak. As a powerful tool, CT provides chest scans in short time. In this study, we presented a deep-learning method for automatic diagnosis of COVID-19 from chest CT images to assist clinicians and radiologists in combating this pandemic.

Butt et. al. (2020) evaluated two CNN-based 3D classification models in their study. One was based on the relatively traditional ResNet-23 model and the other model was designed based on the first network structure which improved the overall accuracy by concatenating the location-attention mechanism in the full-connected layer. The location-attention mechanism used relative distance-from-edge as the extra weight to learn the relative location information of the patch on the pulmonary image. The relative distance-from-edge is the minimum distance from the mask to the center of the patch divided by the diagonal of the minimum circumscribed rectangle of the pulmonary image. Infection foci located near the pleura are more likely to be identified as COVID-19. A total of 528 CT samples (85.4%) were employed to train and valid models, including 189 samples of patients with COVID-19, 194 samples from patients with CP, and 145 samples from healthy people. The remaining 90 CT sets (14.6%) which were selected from people who have not been included in the training stage were used as the test set, including 30 COVID-19, 30 CP, and 30 healthy cases. The location-attention model achieved an AUC of 0.996 for COVID-19 and Non-COVID-19 cases thoracic CT studies and calculated a sensitivity of 98.2% and a specificity of 92.2%.

Ouyang et. al. (2020) developed a novel attention network to automatically diagnose COVID-19 from the CP in chest CT. In particular, they proposed a new online attention module of 3D class activation mapping (CAM) with a 3D CNN to focus on the infected area of the lungs when making diagnostic decisions. The core idea of CAM (Zhou et. al. 2016, Selvaraju et. al. 2017, Fukui et. al. 2019) is to back-propagate the weights of the fully connected layer to the convolution feature map to generate attention maps and they extended this offline operation into a 3D input scene component that can be trained online. Furthermore, they used a dual-sampling strategy to reduce unbalanced learning. The main idea of size-balanced sampling is to repeatedly sample small COVID-19 infection cases and large infection CP cases during the training process. Normally, each sample in the training data set is sent to the network only once with equal probability within one epoch. Due to the imbalance of the infection size distribution, they trained the network through a size-balanced sampling strategy. The purpose was to increase the sampling probability of COVID-19 in small infected areas and CP cases in large infected areas in each mini-batch. Their method was evaluated a large multi-center COVID-19 CT data from eight hospitals. In the training and validation stage, they collected 2186 CT scans from 1588 patients and performed 5-fold cross-validation. In the testing stage, they used another independent large-scale testing data, including 2796 CT scans from 2057 patients. The results showed that their algorithm was able to differentiate COVID-19 from other two classes (CP and normal controls) with 87.5% accuracy, 86.9% sensitivity, 90.1% specificity, 82.0% F1-sorce and 0.944 AUC.

Zhang et. al. (2020) developed an AI system that was able to diagnose COVID-19 and differentiated it from other CP and normal controls. In particular, their AI system identified important clinical markers related to the nature of COVID-19 lesions. Combined with clinical data, their artificial intelligence system could provide accurate clinical prognosis and help clinicians consider appropriate early clinical management and rational allocation of resources. A large CT dataset was constructed using 532,506 CT images from CC-CCII. The COVID-19 diagnosis system was composed of two models: lung lesion segmentation model and diagnosis prediction model. They first trained a segmentation network, using 4,695 COVID-19 and manual segmentation images of CP patients. Then, they trained a classification model using 361,221 CT images from 2,246 patients including 752 COVID-19 patients, 797 CP patients, and 697 normal control patients. The diagnostic classifier took the previous lung lesion map as input, and used the classification network to generate the three-level probabilities of COVID-19, CP and normal controls and produce the class with the maximum probability. The structure of the 3D classification network was adapted from 3D ResNet-18. The network employed multiple 3D convolutional blocks with residual connections to continuously extract local and global context features, and used a fully-connected layer and a softmax activation function to calculate the final prediction. Their model was able to discriminate COVID-19 from CP and normal controls with 92.49% accuracy, 94.93% sensitivity, 91.13% specificity, and an AUROC of 0.9797 on an internal validation dataset. The overall performance for three-way classification obtained an accuracy of 92.49% and an AUROC of 0.9813. In addition, they conducted an AI-assisted clinical prognosis assessment based on CT quantitative parameters and clinical metadata.

In this study, we employed a variety of 3D ResNet models and finally determined the best model as 3D ResNet-18. We proposed a preprocessing method that was to superimpose CT slices into volumes of different depths. We raised the issue of the impact of depth on classification performance and proved that depth 4 had the largest improvement in model performance instead of 64. A total of 110,420 complete CT images (80.4%) were employed to train and valid our model and the remaining 26,836 CT images (19.6%) were used as the test set. Our model has very high performance, achieving recall of 99.96%, precision of 99.35%, f1-sorce of 99.65%, three-way classification accuracy of 99.24% and AUC of 0.9986. We believe that our high performance can be attributed to a large, high-quality dataset we employed and different depths used to train 3D models. Our deep-learning model can alleviate the significant need for diagnostic expertise when the health system is overburdened in pandemic situations or remote areas. Currently, our model is designed to help radiologists and clinicians as an effective first-time screening tool as this can reduce patient waiting time and shorten diagnostic workflow time, thereby lessening the overall workload of radiologists, allowing them to respond quickly and effectively in emergency situations.

## Data Availability

Dataset of the CT images and metadata are constructed from cohorts from the China Consortium of Chest CT Image Investigation (CC-CCII). All CT images are classified into novel coronavirus pneumonia (NCP) due to SARS-CoV-2 virus infection, common pneumonia and normal controls. This dataset is available globally with the aim to assist the clinicians and researchers to combat the COVID-19 pandemic.

http://ncov-ai.big.ac.cn/download?lang=en

